# Artificial Intelligence for Contextual Well-being: Protocol for an Exploratory Sequential Mixed Methods Study with Medical Students as a Social Microcosm

**DOI:** 10.1101/2025.03.09.25323629

**Authors:** Yao Xie, Kayode Philip Fadahunsi, John Broughan, John O’ Donoghue, Joseph Gallagher, Walter Cullen

**Author notes:** Corresponding author: (YX). **Funding** This research received no specific grant from any funding agency in the public, commercial or not-for-profit sectors. **Competing interests** The authors have declared that no competing interests exist. **Data availability** This is the protocol for the upcoming study, so at this point, we do not have any data available to share.

## Abstract

**Introduction:** AI-powered conversational agents have proven effective in alleviating psychological distress, however, concerns about autonomy and authentic psychological development remain, especially in youth during critical stages of identity and resilience formation. Despite the increasing use of AI technologies, there is a significant gap in well-being literacy within educational systems. This gap leaves young adults ill-prepared to navigate the complexities of real world challenges, contributing to rising rates of anxiety, stress, and depression. Furthermore, the lack of AI literacy can exacerbate psychological distress, negatively impacting academic performance and overall well-being. As young adults actively engage with AI, efforts should focus not on resisting technological progress but on fostering their development as users who are capable, aware, and ethical in addressing their contextual well-being needs.

This study aims to extend the understanding of the factors influencing well-being and determine how to harness artificial intelligence for contextual well-being from a human-centred perspective.

**Methods and analysis:** The research is an exploratory sequential mixed methods Study, combining semi-structured interviews and an electronic Delphi study (eDelphi) to gather insights for consensus building. The study adopts a pragmatism paradigm with a foresight approach, ideal for addressing the dynamic, evolving intersection of AI and well-being. Both reflective thematic analysis and descriptive statistical analysis will be used. Medical students (aged 18–30) were selected as a social microcosm study cohort representing youth.

**Ethics and dissemination:** This study was approved by the Human Research Ethics Committee (HREC); Reference: LS-C-24-375-Xie-Cullen.The outcomes of the study will be communicated through publications in peer-reviewed journals, presentations at academic conferences.

## Introduction

Globally, one in four individuals will develop a mental health disorder in their lifetime(1). The COVID-19 pandemic has exacerbated this crisis, driving sharp increases in anxiety, depression, and burnout worldwide(2, 3). Notably, among those most vulnerable are youth aged 15–29, a demographic in which suicide ranks as the third leading cause of death(4). Students, in particular, face compounded risks due to intersecting stressors such as academic pressure, digital hyperconnectivity, and the destabilising transition to adulthood(5). Critically, these challenges are hard to navigate with insufficient intrinsic (e.g., resilience) and extrinsic (e.g., social support) mechanisms, disrupting the equilibrium required to sustain their well-being(5). This disruption is not isolated, well-maintained well-being not only serve as a protective buffer against mental health crises and associated physical consequences(6), but also reflects the profound interconnectedness of these domains, wherein each dynamically influences and reinforces the others(7).

For example, in our previous study(8), it was found that international health care students, a group often exposed to high-stress environments, face unique challenges due to their demanding training and adaptation to new cultural and professional contexts(9). Despite these pressures, well-being support for youth remains inadequate or neglected(10), with insufficient resources to address their diverse identity needs including social, biological, cultural, spiritual, professional and self. This lack of support perpetuates existing inequalities and creates gaps in access to opportunities, resources, and support across various domains, such as education, workplaces, and social interactions and even within families. An individual’s well-being is deeply tied to the contexts they navigate, a phenomenon also referred to as contextual well-being(11). Contextual well-being, grounded in socio-ecological systems(12), acts as a sustainable buffer against mental and physical health issues. When mismatched (e.g., inadequate support for cultural adaptation, overloaded responsibilities), it fractures, exposing individuals to cascading mental and physical health consequences (13, 14).

Our findings indicate that contextual well-being is dynamic, shaped by the continuous interaction between individuals and their environments. It emerges from the interplay between an individual’s identities, the resources and responsibilities associated with their dynamic roles, and the external demands of their environment. For young adults, this includes academic settings, social relationships, professional training environments and beyond, and their roles changes across contexts(15). Addressing contextual well-being not only requires tailored interventions that consider these diverse and evolving needs as external support, but also well-being literacy as the core resources for internal facilitation.

This interplay becomes even more critical as humanity is entering a new era shaped by rapid advancements in artificial intelligence (AI) technologies. For the first time, AI is restructuring the very environments that shape well-being, particularly for youth growing up in digitally saturated societies (16, 17). AI’s integration into education, healthcare, and social interaction redefines developmental contexts, altering how young people access resources, form identities, and manage external demands (16, 17).

Adding to these difficulties, because AI can affect users both favourably and unfavourably, it is crucial to research it in relation to wellbeing(18, 19). AI-empowered conversational agents (AI-CAs) have been defined(20) as “the study of techniques for creating software agents that can engage in natural conversational interactions with humans”(21) and as “software systems that mimic interactions with real people”(22). AI-CAs have proven effective in easing psychological distress and promoting well-being, making mental health support more accessible and widely scalable(17, 20, 23). However, the adoption of AI-CAs also presents risks in some contexts, such as exacerbating job stress and burnout, highlighting the need for careful implementation(24). These underscore the need for a human-centred perspective that prioritises the ethical, equitable, and context-specific implementation of AI for genuine human needs.

Current AI-CAs provide personalised support by understanding and responding to individuals’ contextual needs, reflecting the core idea of contextual well-being. For example, studies show that AI-CAs demonstrate a disposition and capacity for empathy and emotional support(25), facilitate learning, are easy to use and access, and assist users in taking responsibility for their actions, improve users’ engagement and reduce strain on healthcare resources(25). All of which contribute to generally favourable assessments of usability and satisfaction outcomes in healthcare settings(25). However, this same capability raises fundamental concerns about autonomy and authentic psychological development(26), particularly among young people, during the formative years when identity and resilience are being shaped. Additionally, its inherent limitation, the “black box,” which obscures how complex AI language models make decisions, makes it challenging to monitor the advice provided to users(27). Additionally, cost-effectiveness, privacy, system integration, and AI systems design are other major concerns(25). A systematic review(28) found that many AI systems overlook key socio-environmental factors like institutional pressures and cultural norms, reducing their effectiveness in successful well-being related intervention and leading to bias and low user engagement.

Concurrently, youths confront dual critical deficits, i) youths are ill-equipped with the necessary skills as resources to sustain well-being as a condition of equilibrium or balance(29), which later leads to either mental or physical disorders. There is a substantial gap in well-being literacy within educational institutions(30); ii) Alongside limited AI literacy, gaps in well-being literacy can lead to increased psychological distress among students due to ineffective use of AI or reliance on inappropriate AI advice for decision-making(31). Without the skills to recognise and address well-being concerns, such as, mental health issues, and make good choice in decision making process which can adversely affect youths’ overall well-being(19). Notably, regardless of education systems’ readiness, young people are actively utilising these technologies across various contexts. This suggests that rather than attempting to resist technological advancement, efforts should focus on guiding its development to serve genuine human needs and cultivate capable and ethical “users” in response to emerging contexts(32). This requires the education system to close the gap, as education is the foundation of social hope and capital, a space where individuals can foster positivity through active participation in educational growth(11).

This study aims to investigate how AI can enhance contextual well-being from a human-centred perspective and to understand the factors influencing youth well-being, with a focus on harnessing AI’s full potential while safeguarding fundamental human values.

Among the youth adults, medical students represent a social microcosm of young adults. They not only reflect broader youth issues, but also college students, and future healthcare professionals. The decline of mental health among youth, particularly those in high-stakes professions like medicine, represents a global public health emergency(33, 34). Medical students are required to rapidly adjust to demanding situations while enduring uniquely intense psychosocial stressors, including competitive training environments, ethical dilemmas in patient care, and prolonged exposure to human suffering (33, 34). Their well-being affects not only their personal health but also the future quality of healthcare, as burnout is linked to reduced clinical empathy and professional attrition (35). Medical students’ experiences offer broad insights into youth well-being, as their stressors overlap with those of other high-pressure cohorts, making them an ideal group for addressing the study’s objectives.

### Research Questions

1. How do medical students’ perceived gaps in well-being literacy influence their ability to sustain well-being?
2. How does contextual AI impact contextual well-being?
3. How does AI-assisted well-being literacy integrating an understanding of AI’s applications, limitations, and ethical considerations with well-being literacy, enable (a) fostering positive behaviours for well-being, (b) realising AI’s potential in well-being interventions, and (c) mitigating associated risks?

## Methods

### Study design

This body of research employs an exploratory sequential mixed methods design, integrating in-depth semi-structured interviews followed by an electronic Delphi (eDelphi) method. As Creswell and Plano Clark (29) emphasise, mixed methods harness diverse perspectives to generate insights transcending standalone qualitative or quantitative approaches. This methodology enables a dual focus: validating prior findings while exploring underexamined intersections between AI and well-being.

Through prioritising lived experiences of medical students (via interviews) before quantitative consensus-building (via eDelphi), the design circumvents “techno-solutionism”—the tendency to prioritise AI’s technical capabilities over human needs. This approach addresses a critical gap in human-AI interaction research, the demanding of frameworks that centre human-cantered perspectives in AI well-being interventions(36-38)

The chosen methods align with the underlying pragmatism paradigm, and coupled with WHO’s foresight approach(39). The WHO commonly uses the foresight approach for strategic planning and policy-making, exploring trends, advancements, and future scenarios(19). It incorporates horizon scanning and futures-thinking to anticipate change and maximise the potential of emerging technologies to address evolving challenges in complex, diverse environments(39). This approach would help this study remain flexible in response to unexpected contexts including new AI technologies breakthrough, or other relevant unforeseen events that may impact on this study.

Pragmatism provides flexibility in choosing and adapting methods, ensuring that the research process remains practical and responsive(40), which is especially suited to the study, which involves real-world, rapidly evolving AI technologies.

Interview instruction has been piloted with a research team member (YX and JB). As is typical of most mixed-methods research, the planned study design is expected to lie somewhere between fixed and emergent designs due to the nature of mixed methods (41). This indicates that there are endpoints along a continuum rather than a distinct dichotomy(41). As a result, the interpretation of the findings from the in-depth interview phase may reshape the specifics of the next stage(41). Overall, each phase builds upon the previous one, producing findings that are both contextually grounded and generalisable to wider settings. Each phase of study will be piloted prior to the commence of the data collection.

Data will be gathered at two phases: through in-depth interviews and the eDelphi, as outlined in Fig 1.

**Fig 1.**
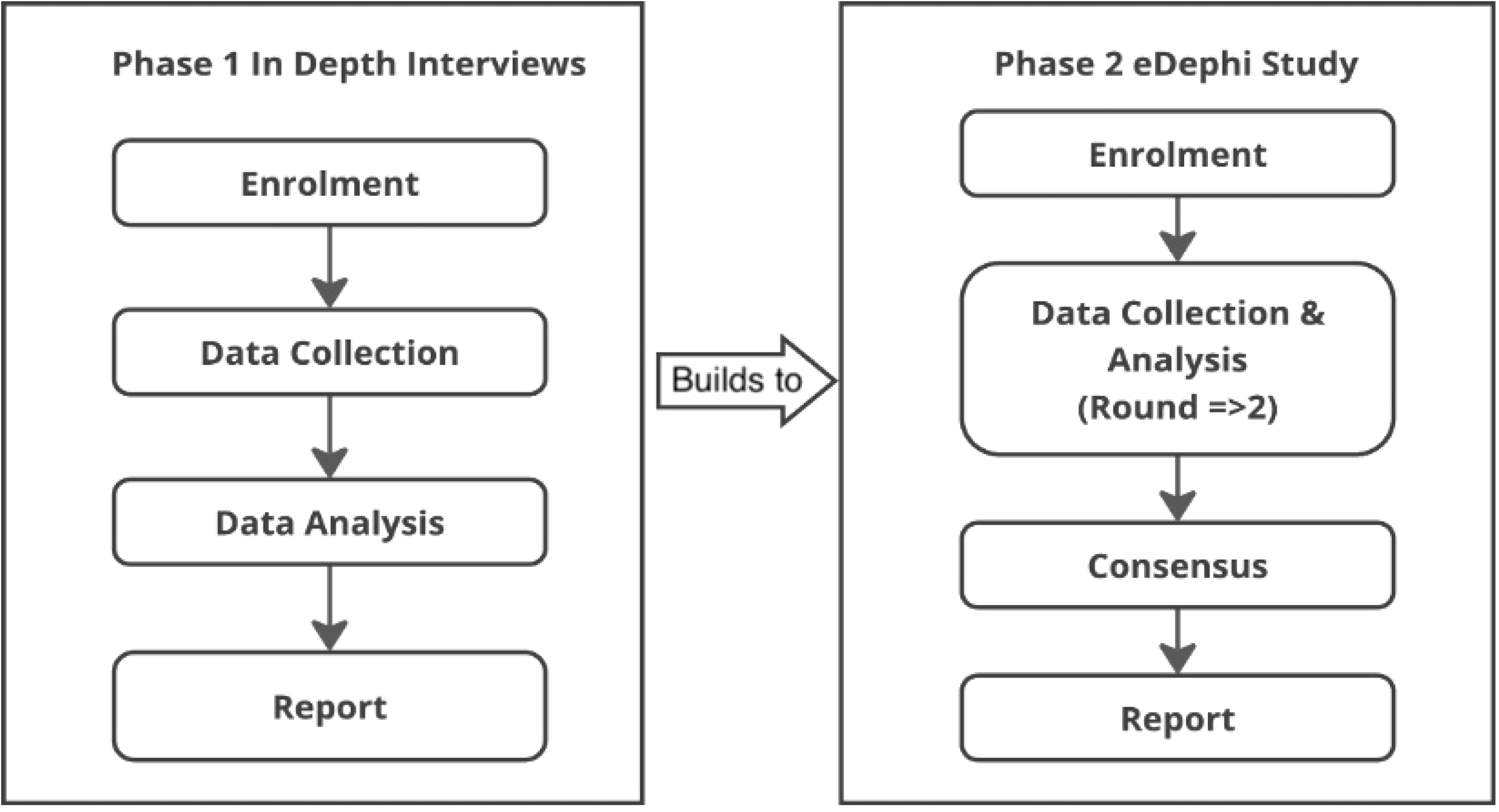
Study procedure

### Sample Size

For phase 1, interviews will be conducted with a target sample of 12 to 20 medical students in higher education, aged 18 to 30 years old. The sample aims for a balanced representation, with approximately half being local students and the other half international students.

This range is considered appropriate for an in-depth exploration of the research objectives(42). While a sample size of 20 participants is generally considered sufficient for qualitative studies (43), saturation, where no new patterns emerge, is typically reached with 9 to 17 interviews(42). Building on this foundation, the study emphasises “information power”, focusing on the richness and depth of the data rather than solely on the number of participants(44). Semi-structured interviews are anticipated to last 60 - 90 minutes, allowing for the generation of quality data that go beyond “surface-level responses”(44).

Delphi studies vary in sample size, with expert panels typically ranging from 4 to 171 participants(45). However, research suggests that a panel of at least 12 participants is typically adequate to reach meaningful consensus(46), with a range of 12 to 50 experts being ideal for obtaining reliable results(47). Beyond a certain threshold, increasing the sample size may result in diminishing returns, with additional input having a reduced impact on the validity of the findings and consensus-building(46, 47). Therefore, in phase 2, the eDelphi study will aim for a sample size between 12 and 20 experts in health and well-being. The emphasis will be on the expertise and relevance of the participants rather than their sheer number. As smaller, highly knowledgeable groups are often more effective in achieving consensus than larger groups with less specialised knowledge, the effectiveness of the eDelphi process hinges more on group dynamics and consensus-building than on statistical power(46).

### Recruitment

Participants will be selected using purposive and snowball sampling methods. Recruitment efforts will utilise internal channels at University College Dublin (UCD), such as the school’s newsletter, and external platforms including LinkedIn, X (formerly Twitter), and other pertinent social media, as well as word of mouth, posters and flyers in relevant areas. Individuals expressing interest will be encouraged to disseminate study-related content on their personal social media accounts to broaden the participant pool. Those who complete interviews will be invited to participate in the subsequent eDelphi phase.

Before scheduling, the participants will receive an information sheet detailing the study’s objectives, confidentiality measures, and their right to withdraw at any time without repercussions. Informed consent will be obtained through an online survey, where participants will be required to read an information sheet and explicitly agree to participate by selecting a consent checkbox before proceeding. This constitutes written electronic consent. The survey will be advertised via QR codes and links on posters and flyers.

All recruitment materials will provide a comprehensive explanation of the study’s objectives, participant selection criteria, and the specifics of involvement. Informed consent will be obtained from all participants to ensure they fully understand the study’s purpose and their role.

### Inclusion and exclusion criteria

For the interviews, medical students aged 18 to 30 will be included in the study as this age range is generally classified as young adults(48). Exclusion criteria include individuals who are not medical students, those under 18 years old, and those over 30 years of age.

For the eDelphi phase, experts from the fields of health and well-being who are interested in contributing to AI for well-being, will be invited to participate as panellists including medical education experts, well-being and mental health professionals, allied health care professionals, and medical students.

## Data Collection

### Phase 1 In-depth Interviews

A semi-structured interview method will be used to explore participants’ perceptions, experiences, and identify research gaps with regards to well-being and AI(49-51). This method balances structure with the flexibility to allow participants to express their thoughts freely(49-51). The is informed by our prior study in this area(8). YX, a PhD researcher with training in qualitative methodologies, will conduct the interviews, which will be held virtually via Zoom for 60–90 minutes. Open-ended questions (see in S1 Appendix) incorporating behavioural event interview techniques will be used. These will encourage participants to share past experiences and behaviours in specific situations, providing context on how they respond to challenges, from awareness and action to the decision-making process for problem-solving(52). Interviews will be audio recorded for transcription and de-identification, with identifiable information replaced by pseudonyms to maintain confidentiality.

### Phase 2 eDelphi method

In phase 2, an eDelphi method will be used to establish consensus among an interdisciplinary panel of healthcare professionals and researchers on how AI could be harnessed to improve well-being for youth. This method is an efficient and cost-effective means to engage participants from geographically diverse locations in Ireland, enhancing inclusivity and avoiding potential power imbalances inherent in in-person discussions (53). The eDelphi process is widely recognised for its structured approach to consensus building(54).

This eDelphi phase will be conducted by an interdisciplinary team with expertise in health, digital health and well-being (YX, KPF, JB, JOD, JG, WC). The study will begin with the development of an open-ended survey questionnaire which will be drafted by YX, drawing on insights from existing literatures and phase one interview data. The survey will then will undergo pilot, collaborative review and iterative refinement by the research team.

The eDelphi survey will be distributed to the expert panel via Qualtrics Responses and will be systematically analysed to identify key themes and concepts, which will then inform the development of structured questionnaires for subsequent rounds. Participants will evaluate and provide feedback on these items, for example, the definition of AI-assisted well-being literacy. The level of agreement will be assessed. Each round of the eDelphi process will span two weeks, with two follow-up reminder emails sent to non-respondents. It is anticipated that the process will consist of 2-3 rounds.

The research team will evaluate participants’ feedback to determine whether survey items should be modified, retained, or removed, in accordance with predefined criteria and stopping rules. For this study, we anticipate a higher consensus threshold of 70% agreement among experts will be applied to identify agreement on survey items, as opposed to the 60% threshold commonly reported in prior research(53).

## Data Analysis

To ensure anonymity, pseudonyms will be used, and any information that could reveal the participants’ identities will be removed.

In Phase 1, interview transcripts will be analysed using NVivo software. Braun and Clarke’s reflective thematic analysis method will be used to identify, analyse, and interpret recurring themes and insights within participants’ responses(55). YX will code the transcripts, the initial codes to emerging themes will be iteratively reviewed by research team members (JB, JOD, KPF, JG and WC) to ensure analytical rigor.

In Phase 2, the analysis will use a mixed-methods approach across sequential eDelphi rounds. Qualitative data from open-ended responses will be analysed using Braun and Clarke’s reflective thematic analysis (55) to identify common themes and patterns relevant to well-being, and the integration of AI into potential well-being initiatives. Particularly from the initial round, which will inform the development of structured questionnaires for subsequent rounds. Quantitative data, including Likert-scale responses indicating levels of agreement, and participants’ demographic characteristics will be analysed using descriptive statistics (such as means, medians) to assess consensus among experts.

### Timeline

The study was approved by UCD Human Research Ethics Committee (HREC) on January 10, 2025. Phase 1, involving interview recruitment and data collection, is scheduled as follows: recruitment will take place from February 19 to April 30, 2025, with data collection occurring between May 1 and June 30, 2025. Data analysis for Phase 1 is expected to be completed by July 2025. Phase 2, which will involve the eDelphi method, will begin with the recruitment of panellists from July 15 to August 15, 2025. The eDelphi rounds (Rounds 1 and 2) will be conducted between August 20 and October 31, 2025, followed by data analysis, which is projected to conclude in November 2025. The final results are anticipated to be available by January 2026.

## Discussion

This study investigates how AI may improve well-being from a human-centred standpoint and understanding the factors influencing youths’ well-being using medical students as an social microcosm. To the best of our knowledge, this study is the first to hypothesise that contextual AI can empower contextual well-being and introduce the concept of AI-assisted well-being literacy.

An anticipated outcome is that the integration of AI literacy and well-being literacy will equip individuals with the skills to engage with AI responsibly, critically, and with contextual awareness. This includes preserving ethics, moral principles, and decision-making autonomy while ensuring that AI interventions do not undermine cognitive development, especially among youths. The integration of Al-assisted well-being literacy into educational curricula is essential to strike a balance between embracing technological progress and safeguarding collective well-being. AI functions as a powerful behavioural stimulator with a dual influence on human psychology and behaviours(56). Users engage with AI systems across multiple, fluid identity in their contextual well-being. For instance, as students seeking academic guidance, friends navigating social support, or professionals managing workplace demands. Context determines how, when, and why individuals interact with AI. Crucially, not all roles or contexts align equally with AI’s utility: some identities (e.g., mental health patients in crisis) may require cautious or restricted AI use to avoid harm, while others (e.g., learners accessing tutoring) benefit from proactive engagement.

Efforts by multiple stakeholders are driving advancements in language models, and users must be prepared and understand how to leverage AI technologies to support well-being, while maintaining youths’ ability to make informed, ethical decisions across various contexts, These decisions not only shape individual well-being but also influence community and societal well-being. Therefore, Further research is needed to develop comprehensive educational frameworks that merge AI literacy with well-being literacy.

One of the key strengths of this research lies in its methodological design, which combined qualitative interviews and an eDelphi study to ensure depth and breadth in data collection and analysis, each study will informed by another, and both are informed by the evidence from our previous study(8).The semi-structured interviews will allow participants to share detailed, personal perspectives on well-being and AI(49-51), while the eDelphi method will facilitate consensus-building among experts in AI for well-being(54). Another strength is the inclusion of diverse participants, representing various backgrounds and expertise, which enriches the findings’ applicability across different contexts.

However, every study has limitations(57). In this research, and well-being are culturally and context-dependent, meaning the experiences of medical students in one country or region may not reflect those of students in other parts of the world. This is particularly relevant given the diverse ways in which mental health is perceived and addressed across societies. To mitigate this limitation, we have previously conducted an international cohort analysis through a systematic integrative review and will aim to include a diverse demographic in our sample. Given the exploratory nature of the study and the novelty of AI, the research may not fully explore every facet of AI’s impact on well-being, particularly in terms of long-term effects. However, the study provides foundational data that can guide future, more structured, hypothesis-driven research.

## Dissemination

The outcomes of the study will be communicated through publications in peer-reviewed journals, presentations at academic conferences.

## Data Availability

No datasets were generated or analysed during the current study. All relevant data from this study are available from the corresponding author on reasonable request.

## Ethics

This study was approved by the UCD HREC; Reference:LS-C-24-375-Xie-Cullen. Although the proposed study is considered low risk, it is important to acknowledge that all research involving human participants carries some level of risk. Ethical guidelines require the identification, minimisation, and disclosure of potential risks, even in studies with minimal risk.

Confidentiality and privacy will be strictly maintained during recruitment and data collection. Participants will be informed of any risks or benefits of their involvement, with a clear focus on obtaining informed consent. The study will also consider cultural sensitivity and language proficiency. To ensure inclusivity, participants from diverse cultural backgrounds will be respected. Non-native English speakers will be assured that their language proficiency will not be judged, creating a supportive environment for all participants.

Confidential data collected in this study will be securely stored for five years on a password-protected computer, with access limited to authorised personnel only (UCD research team members). Electronic data will be stored in password protected files on encrypted computers belonging to members of the research team. The data will be securely stored on UCD’s cloud-based infrastructure, which includes encrypted servers.

All audio recordings will be transcribed in a way that guarantees complete anonymity, with no identifiable information retained. Upon completion of the transcription, the original recordings will be permanently deleted. Both the transcribed data and any ensuing analyses or publications will exclude personal identifying information, ensuring participant confidentiality is maintained throughout. Notably, the eDelphi process, where responses will be anonymised to prevent bias and ensure honest feedback. All data will be securely stored and handled according to ethical standards.

## Authors’ contributions

YX conceptualised the study, drafted and edited the manuscript, visualisation and followed through the study process as part of her doctoral study. JOD contributed to the selection of the methodology. YX, KPF, JB, JOD, JG, and WC are members of the research team. KPF, JB, JOD, JG, and WC provided expert feedback, reviewed, and edited the manuscript. KPF, JOD, JG, and WC were involved in co-supervision.

## Acknowledgements

YX would show her gratitude to her supervisors and research team’s support on her doctoral project.

## Supporting information

S1 Appendix

## Notes

### Competing Interest Statement

The authors have declared no competing interest.

### Funding Statement

The author(s) received no specific funding for this work.

### Author Declarations

This study was approved by the UCD Human Research Ethics Committee (HREC) Reference:LS-C-24-375-Xie-Cullen.

## References

1. Kessler RC, Berglund P, Demler O, Jin R, Merikangas KR, Walters EE. Lifetime prevalence and age-of-onset distributions of DSM-IV disorders in the National Comorbidity Survey Replication. Arch Gen Psychiatry. 2005;62(6):593–602.

2. Xiong J, Lipsitz O, Nasri F, Lui LMW, Gill H, Phan L, et al. Impact of COVID-19 pandemic on mental health in the general population: A systematic review. J Affect Disord. 2020;277:55–64.

3. World Health Orgnization. COVID-19 pandemic triggers 25% increase in prevalence of anxiety and depression worldwide 2022. [Available from: https://www.who.int/news/item/02-03-2022-covid-19-pandemic-triggers-25-increase-in-prevalence-of-anxiety-and-depression-worldwide?

4. World Health Orgnization. Mental health of adolescents: WHO; 2024 [Available from: https://www.who.int/news-room/fact-sheets/detail/adolescent-mental-health.

5. Zolopa C, Burack JA, O’Connor RM, Corran C, Lai J, Bomfim E, et al. Changes in youth mental health, psychological wellbeing, and substance use during the COVID-19 pandemic: A rapid review. Adolescent Research Review. 2022;7(2):161–77.

6. McEwen BS. Stress, adaptation, and disease: Allostasis and allostatic load. Annals of the New York academy of sciences. 1998;840(1):33–44.

7. Steptoe A, Deaton A, Stone AA. Subjective wellbeing, health, and ageing. Lancet. 2015;385(9968):640–8.

8. Xie Y, Fadahunsi KP, Flynn P, Taylor-Robinson S, Gallagher J, Cullen W, et al. Barriers and Facilitators of International Health Care Students’ Well-Being in Higher Education: Protocol for a Systematic Integrative Review. JMIR Research Protocols. 2024;13(1):e59927–e.

9. Astfalk T, Muller-Hilke B. Same same but different - A qualitative study on the development and maintenance of personal networks among German and international medical students. GMS J Med Educ. 2018;35(5):Doc58.

10. Uhlhaas PJ, Davey CG, Mehta UM, Shah J, Torous J, Allen NB, et al. Towards a youth mental health paradigm: a perspective and roadmap. Mol Psychiatry. 2023;28(8):3171–81.

11. Carter S, Andersen C. Wellbeing in Educational Contexts–Second edition: University of Southern Queensland; 2023.

12. Bronfenbrenner U. The ecology of human development: Experiments by nature and design: Harvard university press; 1979.

13. Masten AS. Global perspectives on resilience in children and youth. Child development. 2014;85(1):6–20.

14. McNulty JK, Fincham FD. Beyond positive psychology? Toward a contextual view of psychological processes and well-being. American Psychologist. 2012;67(2):101.

15. Multisystemic Resilience: Adaptation and Transformation in Contexts of Change: Oxford University Press; 2021. Available from: 10.1093/oso/9780190095888.001.0001

16. Bundy A, Chater N, Muggleton S. Introduction to ‘Cognitive artificial intelligence’. Philos Trans A Math Phys Eng Sci. 2023;381(2251):20220051.

17. Balan R, Dobrean A, Poetar CR. Use of automated conversational agents in improving young population mental health: a scoping review. npj Digital Medicine. 2024;7(1):75.

18. Ettman CK, Galea S. The Potential Influence of AI on Population Mental Health. JMIR Ment Health. 2023;10:e49936.

19. Shahzad MF, Xu S, Lim WM, Yang X, Khan QR. Artificial intelligence and social media on academic performance and mental well-being: Student perceptions of positive impact in the age of smart learning. Heliyon. 2024;10(8):e29523.

20. Mariani MM, Hashemi N, Wirtz J. Artificial intelligence empowered conversational agents: A systematic literature review and research agenda. Journal of Business Research. 2023;161:113838.

21. Khatri C, Venkatesh A, Hedayatnia B, Gabriel R, Ram A, Prasad R. Alexa prize—state of the art in conversational AI. AI magazine. 2018;39(3):40–55.

22. Radziwill NM, Benton MC. Evaluating quality of chatbots and intelligent conversational agents. arXiv preprint arXiv:170404579. 2017.

23. Li H, Zhang R, Lee Y-C, Kraut RE, Mohr DC. Systematic review and meta-analysis of AI-based conversational agents for promoting mental health and well-being. npj Digital Medicine. 2023;6(1):236.

24. Kim B-J, Lee J. The mental health implications of artificial intelligence adoption: the crucial role of self-efficacy. Humanities and Social Sciences Communications. 2024;11(1):1561.

25. Milne-Ives M, de Cock C, Lim E, Shehadeh MH, de Pennington N, Mole G, et al. The Effectiveness of Artificial Intelligence Conversational Agents in Health Care: Systematic Review. J Med Internet Res. 2020;22(10):e20346.

26. Ahmad SF, Han H, Alam MM, Rehmat MK, Irshad M, Arraño-Muñoz M, et al. Impact of artificial intelligence on human loss in decision making, laziness and safety in education. Humanities and Social Sciences Communications. 2023;10(1):311.

27. Duran JM, Jongsma KR. Who is afraid of black box algorithms? On the epistemological and ethical basis of trust in medical AI. J Med Ethics. 2021.

28. Malgaroli M, Hull TD, Zech JM, Althoff T. Natural language processing for mental health interventions: a systematic review and research framework. Translational Psychiatry. 2023;13(1):309.

29. Dodge R, Daly AP, Huyton J, Sanders L. The challenges of defining wellbeing. International journal of wellbeing. 2012;2(3):222–35.

30. Oades LG. Wellbeing Literacy: The Missing Link in Positive Education. In: White MA, Slemp GR, Murray AS, editors. Future Directions in Well-Being: Education, Organizations and Policy. Cham: Springer International Publishing; 2017. p. 169–73.

31. Shen Y, Cui W. Perceived support and AI literacy: the mediating role of psychological needs satisfaction. Front Psychol. 2024;15:1415248.

32. Nguyen A, Ngo HN, Hong Y, Dang B, Nguyen BT. Ethical principles for artificial intelligence in education. Educ Inf Technol (Dordr). 2023;28(4):4221–41.

33. Dyrbye LN, Thomas MR, Massie FS, Power DV, Eacker A, Harper W, et al. Burnout and suicidal ideation among U.S. medical students. Ann Intern Med. 2008;149(5):334–41.

34. Quek TT, Tam WW, Tran BX, Zhang M, Zhang Z, Ho CS, et al. The Global Prevalence of Anxiety Among Medical Students: A Meta-Analysis. Int J Environ Res Public Health. 2019;16(15).

35. Ligibel JA, Goularte N, Berliner JI, Bird SB, Brazeau CMLR, Rowe SG, et al. Well-being parameters and intention to leave current institution among academic physicians. JAMA Network Open. 2023;6(12):e2347894–e.

36. World Health O. Ethics and governance of artificial intelligence for health: large multi-modal models. WHO guidance: World Health Organization; 2024.

37. Kasinidou M, Kleanthous S, Barlas P, Otterbacher J, editors. I agree with the decision, but they didn’t deserve this: Future developers’ perception of fairness in algorithmic decisions 2021.

38. Floridi L, Cowls J. A unified framework of five principles for AI in society. Machine learning and the city: Applications in architecture and urban design. 2022:535–45.

39. World Health O. Foresight approaches in global public health: a practical guide for WHO staff: World Health Organization; 2022.

40. Kaushik V, Walsh CA. Pragmatism as a Research Paradigm and Its Implications for Social Work Research. Social Sciences [Internet]. 2019; 8(9).

41. Creswell JW, Clark VLP. Designing and conducting mixed methods research: Sage publications; 2017.

42. Hennink M, Kaiser BN. Sample sizes for saturation in qualitative research: A systematic review of empirical tests. Soc Sci Med. 2022;292:114523.

43. Frechette J, Bitzas V, Aubry M, Kilpatrick K, Lavoie-Tremblay M. Capturing lived experience: Methodological considerations for interpretive phenomenological inquiry. International Journal of Qualitative Methods. 2020;19:1609406920907254.

44. Flynn O, Blake C, Fullen BM. A qualitative exploration of migraine in students attending Irish Universities. Plos one. 2024;19(8):e0305643.

45. Skulmoski GJ, Hartman FT, Krahn J. The Delphi method for graduate research. Journal of Information Technology Education: Research. 2007;6(1):1–21.

46. Vogel C, Zwolinsky S, Griffiths C, Hobbs M, Henderson E, Wilkins E. A Delphi study to build consensus on the definition and use of big data in obesity research. International Journal of Obesity. 2019;43(12):2573–86.

47. Staykova MP. Rediscovering the Delphi technique: a review of the literature. Advances in Social Sciences Research Journal. 2019;6(1).

48. In: Bonnie RJ, Stroud C, Breiner H, editors. Investing in the Health and Well-Being of Young Adults. Washington (DC) 2015.

49. Nyanchoka L, Tudur-Smith C, Porcher R, Hren D. Key stakeholders’ perspectives and experiences with defining, identifying and displaying gaps in health research: a qualitative study protocol. BMJ Open. 2019;9(8):e027926.

50. Adeoye‐Olatunde OA, Olenik NL. Research and scholarly methods: Semi‐structured interviews. Journal of the american college of clinical pharmacy. 2021;4(10):1358–67.

51. Britten N. Qualitative interviews in medical research. BMJ. 1995;311(6999):251–3.

52. Ravindranath S. Behavioral Event Interview. Qualitative Techniques for Workplace Data Analysis: IGI Global; 2019. p. 73–95.

53. Fadahunsi KP, Wark PA, Mastellos N, Gallagher J, Majeed A, Car J. Clinical information quality of digital health technologies: protocol for an international eDelphi study. BMJ open. 2022;12(4):e057430.

54. Barrett D, Heale R. What are Delphi studies? Evidence Based Nursing. 2020;23(3):68.

55. Byrne D. A worked example of Braun and Clarke’s approach to reflexive thematic analysis. Quality and Quantity. 2022;56(3):1391–412.

56. Sadeh-Sharvit S, Camp TD, Horton SE, Hefner JD, Berry JM, Grossman E, et al. Effects of an Artificial Intelligence Platform for Behavioral Interventions on Depression and Anxiety Symptoms: Randomized Clinical Trial. J Med Internet Res. 2023;25:e46781.

57. Ross PT, Bibler Zaidi NL. Limited by our limitations. Perspect Med Educ. 2019;8(4):261–4.

